# Handheld, quantitative phase imaging probe for skin evaluation at subcellular resolution *in vivo*, in real-time, and in 3D

**DOI:** 10.64898/2026.11.10.26367486

**Authors:** Srinidhi Bharadwaj, Zhe Guang, Lukas A. Clark, Jacob P. Callaway, Zhenmin Li, Christopher L. Suhr, Samuel N. Lucas, Caroline Serafini, Jimena Garcia-Nuno, Zachary Wolner, Mohammad K. Khan, Susan Napier Thomas, Francisco E. Robles

**Author notes:** These authors contributed equally to this work (Lukas A. Clark, Jacob P. Callaway).

## Abstract

Skin cancer is the most common malignancy in the United States. The gold standard for diagnosis of skin cancer is histopathological analysis via tissue biopsy. However, identifying which lesions to biopsy remains a significant clinical challenge, particularly at early stages of disease. In this work, we present a volumetric quantitative oblique back-illumination microscopy (qOBM) probe, a highly maneuverable, handheld imaging system capable of providing high-resolution, 3D visualization of the skin microstructure. We show the probe is capable of visualizing the dynamic, layered microstructure of the epidermis and the top of the dermis across all Fitzpatrick skin types in human skin *in vivo*. The easy maneuverability of the probe is highlighted by imaging anatomically challenging areas typically hard to reach for bulkier systems. Finally, we present additional case studies in *ex vivo* murine models and *in vivo* human nevi that further illustrate the potential clinical applications of the volumetric handheld probe. These results establish a low-cost, easy to use, high-resolution, compact system that combines the strengths of its predecessors – the excellent maneuverability of the widely-used dermatoscopes and 3D, high-resolution imaging of prior bulkier systems – to improve screening of dermatological lesions at the bedside.

## 1. INTRODUCTION

The skin covers nearly the entire external surface of the body, forming an initial line of defense against environmental pathogens and threats, such as UV exposure. Yet, skin cancer is the most common malignancy in the United States even though many cases are highly curable through early risk-reduction practices, such as monitoring of the tissue surface.^1^ The three most common skin cancers – basal cell carcinoma, squamous cell carcinoma, and melanoma – arise from the most superficial layer of the skin, the epidermis. The keratinocytes of the epidermis are the first cells impacted by UV exposure, and melanocytes are primarily located in the basal layer of the epidermis.^2^

For skin cancer, lower mortality and morbidity rates are strongly associated with early detection and diagnosis.^1^ Early detection screenings, such as self-skin examinations and dermatological screenings, are routinely used to aid in early diagnosis and subsequent treatment,^3^ but evidence indicates that visual inspection of lesions yields inconclusive results.^4,5^ Lesions on pigmented skin can also be more difficult to identify and are associated with greater morbidity and mortality.^6^ The gold standard of histopathological analysis requires a biopsy and can take several days to yield actionable results.^1^ However, biopsies have several limitations, including partial sampling of a lesion, scarring from the biopsy procedure, and discomfort during the biopsy procedure. Critically, biopsies are also subject to the initial equivocal visual assessment which is again inconclusive and can lead to missing cancerous lesions or unnecessary biopsy of normal nevi. Moreover, if patients present with many concerning lesions, not all lesions can be biopsied. Patients may also have suspicious lesions in sensitive areas, such as the face or genitalia, where biopsy has more significant repercussions.

To aid the dermatologist in this initial selection of lesions for biopsy, various imaging technologies have been developed. Dermatoscopy is the most commonplace method for imaging skin lesions in the dermatology clinic.^7^ Handheld dermatoscopes are compact, easy-to-use devices that provide a clinician with a 10x magnified image of the surface of a lesion of interest. As handheld, polarized magnifying glasses, dermatoscopes have excellent maneuverability; any skin lesion of concern over the whole body can be easily assessed by a dermatologist with a dermatoscope. However, since dermatoscopy cannot provide images with cellular resolution, dermatologists must be trained to correlate features found on dermatoscopy with histopathological features. In addition, dermatoscopes only provide a singular 2D plane image of the lesion; thus, dermatoscopy cannot be used to probe the complex, dynamic three-dimensional microstructure of the skin. Hence, despite its widespread usage, the diagnostic utility of dermatoscopy in determining cancerous lesions is equivocal and highly dependent on the expertise of the user.^7,8^

Imaging modalities that provide 3D structure and higher spatial resolution have been developed, including confocal reflectance microscopy (RCM) and optical coherence tomography (OCT). RCM is an optical imaging technique that provides cellular, volumetric resolution of the skin and has moderate integration in dermatology clinics.^9^ Like many optical techniques, RCM is limited by a penetration depth of ∼200 µm into the tissue. OCT is another optical imaging technique that provides volumetric cellular resolution and improved penetration depth in thick tissue.^10^ While high-resolution, conventional OCT methods have low cellular contrast. Line-field confocal OCT (LC-OCT) was developed to address this limitation and is a technique that provides cellular resolution of skin to a depth of ∼0.5 mm into the tissue.^11^ However, for both RCM and OCT, high-resolution imaging in 3D can come at the cost of maneuverability. Widespread adoption is limited by cost, complexity, and bulkiness of the systems.^12^ These limitations elucidate a need for system that can operate at the best of both worlds – a high-resolution, 3D imaging technique that is compact, low-cost and easy to use such that it can be maneuvered to image anywhere on the skin.

We recently introduced quantitative oblique back-illumination microscopy (qOBM), an optical imaging technique that provides label-free, non-invasive, and real-time quantitative phase information in thick, highly scattering tissues.^13,14^ Like quantitative phase imaging (QPI), qOBM provides cellular and subcellular contrast with nanometer scale sensitivity based on the intrinsic refractive index properties of the sample.^15,16^ A critical difference, however, is that qOBM can provide 3D information of thick scattering samples using epi-illumination, while traditional QPI methods cannot. We have used qOBM to image a number of clinical samples with clear cellular and subcellular contrast, including blood, human and rat brain, and a range of organoids.^17–21^ Moreover, qOBM enables 3D imaging of complex biological structure^14^ and has been recently adapted into a handheld probe with a static focal plane designed for the use of imaging brain tumor margins in the operating room, among other applicatoins.^16^

In this work, we extend the utility of the handheld probe by adding critical 3D volumetric imaging capabilities. The result is a uniquely maneuverable microscope that can acquire volumes of the skin that reflect its dynamic microstructure in real-time. Thus, this work combines the strengths of its predecessors – the excellent maneuverability of the widely-used dermatoscopes and 3D, high-resolution imaging of prior bulkier systems. By enabling fast, label-free, volumetric imaging in a compact, low-cost, and highly maneuverable form factor, this probe has the potential to bring depth-resolved visualization of skin microstructure closer to routine clinical practice. Ultimately, this synergistic innovation could provide dermatologists with real-time diagnostic information at the bedside for all skin tones.

## 2. RESULTS

### 2.1. qOBM volumetric probe setup and design

The design of the handheld, volumetric qOBM probe is shown in **Fig. 1A**. The basic design mirrors that of a recently developed handheld qOBM configuration,^16^ but contains key alterations (described below). This probe is an epi-illumination system that shines light from 850 nm LEDs into the skin using 0.75-mm polymethyl methacrylate (PMMA) fibers, which are oriented 90° from each other around an objective (see **Fig. 1A** upper inset). Light is delivered sequentially from the four fibers. Via multiple scattering, some photons are redirected, from the source into the tissue and back towards the tissue surface and into the objective, thus creating a virtual light source within the sample with an oblique illumination, a phenomenon known as oblique back-illumination (see **Fig. 1A** lower inset).^22^ This process is repeated sequentially for each of the four LEDs (coupled to four illumination fibers) to create four oblique back-illumination raw captures. Captures from opposing illumination angles are subtracted to create two orthogonal differential phase contrast (DPC) images.^22,23^ The two DPC images are further deconvolved using a measured transfer function to create a single qOBM image that provides quantitative structural information of the skin (**Fig. 1B-C**).^13,14,24^ Additional verification of the selected fiber lateral separation from the objective and angle of illumination can be found in the **Supplement: Section 2**.

**Figure 1.**
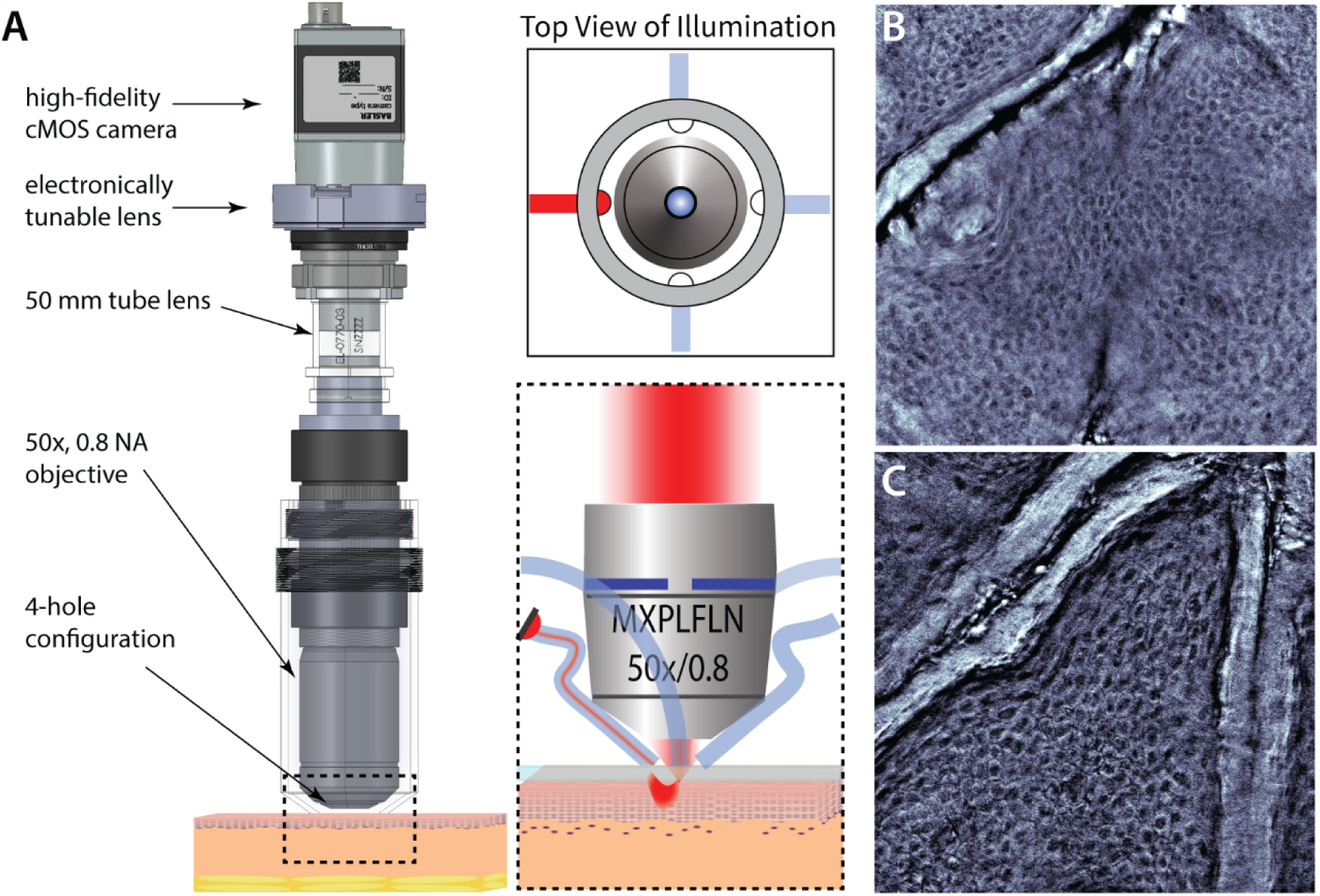
Volumetric qOBM probe design. The sample is illuminated sequentially by four illumination fibers (**A**, insets) and an image is relayed through an Olympus MXPLFLN 50x 0.8 NA objective, a tube lens, and an electronically tunable lens, onto a cMOS camera (**A**). Four captures, one from each angle, are then processed in real-time to create qOBM images of skin (**B**,**C**).

We have simplified the detection optics for this new iteration of the handheld probe by using a single objective (Olympus MXPLFLN 50x 0.8NA) as an initial relay of the image instead of a two objective configuration used previously that includes a finite high-NA gradient refractive index objective.^16^ The objective used here is simply coupled to a 50-mm doublet tube lens (Thorlabs), projecting the image onto a complementary metal-oxide semiconductor (cMOS) camera (Basler ace acA1920-150um Monochrome USB 3.0).

To enable 3D imaging, we add an electronically tunable lens (EL-16-40-TC-VIS-20D) behind the tube lens. This allows us to tune the overall focusing power of the compound lens system, which in turn projects different focal planes onto the detector and achieves 3D imaging with a simple widefield configuration without moving components. The depth range for the volumetric probe is dependent on precise placement of the ETL along the optical axis; the depth range enabled by the ETL in tissue is 436 µm. A theoretical derivation and experimental corroboration of the volumetric depth range of the probe in two configurations can be found in the **Supplement: Section 1**, where additional images of the handheld probe in physical implementation can also be found.

The maximum imaging rate of the volumetric, handheld qOBM probe is 30 frames per second (fps), which is a quarter of the maximum imaging rate of camera used. At this rate, the full field of view (350 µm x 350 µm image) is captured in 33 milliseconds (ms). The settling time of the ETL is 40 ms when traversing half its diopter range (∼200 µm)^25^, so the settling time in single micron adjustments of the ETL are negligible. A single 350 µm x 350 µm x 30 µm volume (with single-micron axial step sizes) can then be acquired in 1 second. The lateral and axial resolution are ∼0.7 µm and 3 µm, respectively, and the working distance of the objective is 3 mm.

Several key advancements have been made since the development of our first-generation probe.^16^ First, this configuration is simpler to align as it does not require precise positioning of the GRIN lens. Crucially, it enables the smooth integration of 3D imaging capabilities without moving components. Lastly, it has a larger working distance and avoids field curvature aberrations previously present due to the GRIN lens. These improved imaging capabilities are critical for dermatological applications.

### 2.2. The volumetric, handheld qOBM probe broadly surveys healthy human skin in vivo

We volumetrically imaged the full thickness of a healthy dorsal hand epidermis using the handheld probe, as shown in **Figure 2**. The most superficial layer of the epidermis, the stratum corneum (**Fig. 2A**), is marked by keratin deposits and prominent, magnified lines of skin microrelief. The next two cellular layers of the epidermis, the stratum granulosum (**Fig. 2B**) and the stratum spinosum (**Fig. 2C**), are marked by an organized network of keratinocytes. Transition from the stratum granulosum to the stratum spinosum can be difficult to observe, but the younger cells of the stratum spinosum are smaller (10-15 µm) than the cells in the stratum granulosum (15-20 µm)^26^, as shown in insets of the same size (30 µm x 30 µm) in **Figure 2B-C**.

**Figure 2.**
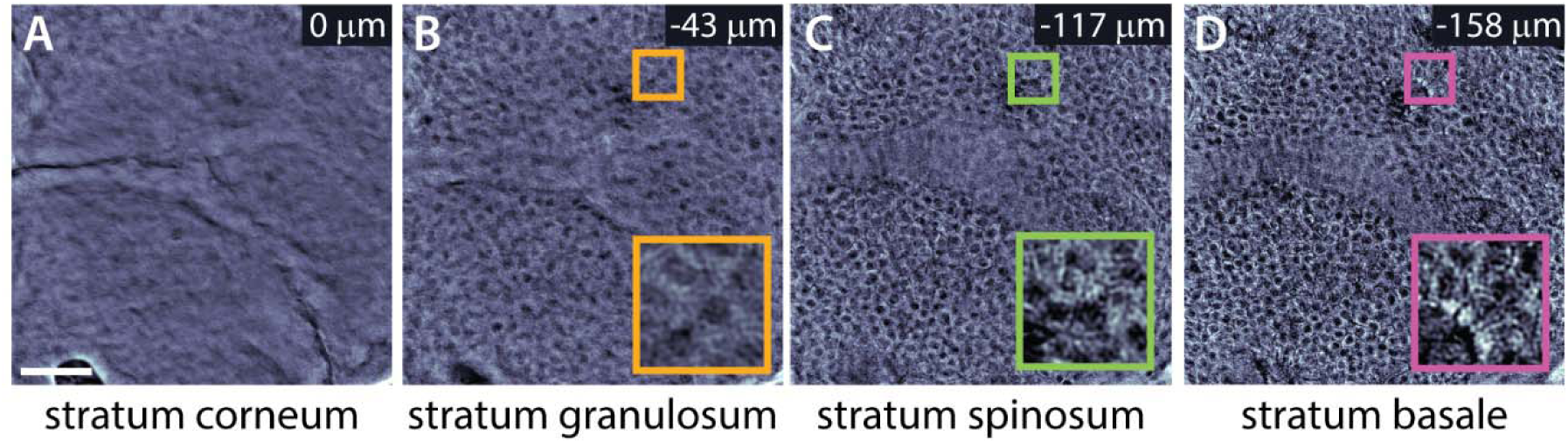
Epidermal layers with the volumetric qOBM probe. For any given skin sample, the volumetric probe visualizes all layers of the epidermis, including the stratum corneum (**A**), stratum granulosum (**B**), stratum spinosum (**C**), and stratum basale (**D**), where blood flow from capillary loops extending from the top of the dermis can be seen (**Supplementary Video 1**). Insets are 30 µm by 30 µm. Scale bar 50 µm.

The deepest layer of the epidermis is the stratum basale, the site of rapid keratinocyte division and hence the smallest keratinocytes (6-10 µm). Capillary blood flow visualization is possible at this layer, as the undulating rete ridges that mark the transition from the epidermis to the dermis are seen (**Fig. 2D**). In healthy skin, melanocytes are concentrated along these rete ridges and can be distinguished from keratinocytes by the bright, highly refractive melanin, as shown in the inset of **Fig. 2D**. The ∼200 µm volume of epidermal thickness shown in **Figure 2** can be fully visualized in 3D in **Supplementary Video 1**, where capillary blood flow can also be visualized in real time. These results highlight the dynamic, three-dimensional microstructure of the epidermis into the dermis that the volumetric probe can extract in real-time.

Epidermal thickness can range from 30-600 µm and varies by site and age.^27,28^ Because skin characteristics are variable across patients, we image subjects across the entire the Fitzpatrick (FP) scale. This scale classifies skin tones into 6 distinct types based on photosensitivity. FP1 is characterized by a high propensity to burn and minimal tanning response; meanwhile, an FP6 is characterized by a low propensity to burn and a robust tanning response.^29^ Generally, higher Fitzpatrick skin tones will have darker skin color. Although this system’s basis in self-reported and subjective classification has its challenges, it is nonetheless a useful tool for confirming the efficacy of our device across a range of skin tones. We used the volumetric handheld probe to image the full epidermis of the dorsal hand across all Fitzpatrick skin types. **Figure 3** shows images from the stratum spinosum/basale layer from the dorsum of the hand of healthy volunteers from each FP type. Across all skin types, we can clearly see “cobblestoning” of the organized network of healthy keratinocytes. Volumetric videos through the full thickness of the epidermis (**Supplementary Video 2**) show the same transitions described in **Figure 2** – from the keratinaceous stratum corneum to the highly cellular stratum basale.

**Figure 3.**
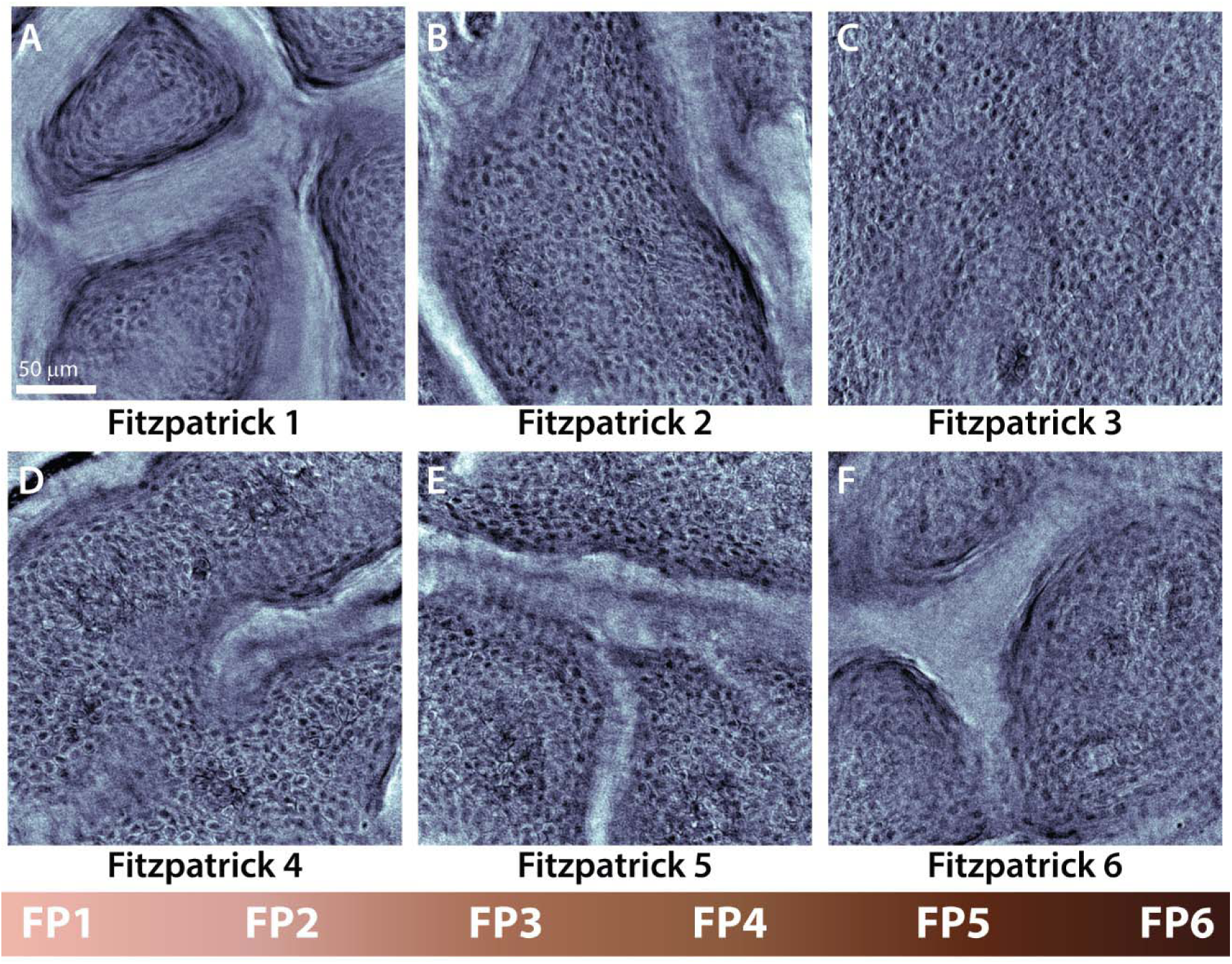
Fitzpatrick skin types with the volumetric qOBM probe. The volumetric skin probe visualizes all Fitzpatrick (FP) skin types, ranging from the lightest skin tone (Fitzpatrick 1, **A**) to the darkest skin tone (Fitzpatrick 6, **F**), shown in A-F at the stratum spinosum layer of the epidermis. Scale bar 50 µm.

Many of these videos also display active capillary blood flow, indicating that the acquisition of real time blood dynamics is possible across all skin types. Notably, the full-thickness volumetric acquisition with blood flow from the FP6 volunteer (**Supplementary Video 2**) demonstrates that light absorption from a relatively higher amount of melanin does not meaningfully affect the image quality, and thus the qOBM probe can achieve subcellular detail across Fitzpatrick skin types. To quantify these observations, we use image entropy at each depth as a measure of information content as performed in prior work.^14,30^ **Supplemental Fig. S4** show the entropy has similar trends throughout the imaged depth for all skin tones and similar entropy values at the dermal-epidermal junction, which again supports the conclusion that qOBM image quality is not significantly affected by skin tone. These results are especially crucial for *in vivo* monitoring of lesions in skin of color with our device, as many lesions can go unrecognized and undiagnosed when they are present in higher FP skin tones.^6^

Facial skin is among the skin’s most chronically sun-exposed regions, making it particularly susceptible to UV-induced damage. Yet, the face’s small, complex contours and anatomically constrained surfaces pose challenges for consistent imaging, especially with bulkier systems. The handheld nature of the qOBM probe and its compact form factor enables *in vivo* imaging in anatomically challenging areas, including the face as shown in **Figure 4**, highlighting the unique maneuverability of a handheld probe. In **Fig. 4A-D**, we show an area depiction (**Fig. 4A**), dermatoscopy (**Fig. 4B**) and qOBM (**Fig. 4C-D**) images of the tip of the ear. We observe clear histological detail in the qOBM images including a potential Langerhans cell in **Figure 4C**, which has similar morphology to a melanocyte as demonstrated in RCM literature.^31^ However, this cell displays a uniform increase in refractive index as opposed to the punctate bright spots of the melanosomes contained within the melanocytes. We also see hair (orange arrow, **Fig. 4D**) and large pockets of sebum or keratin buildup on the ear (green arrow, **Fig. 4D**), a site on the body that demonstrates robust sebum production.^32^

**Figure 4.**
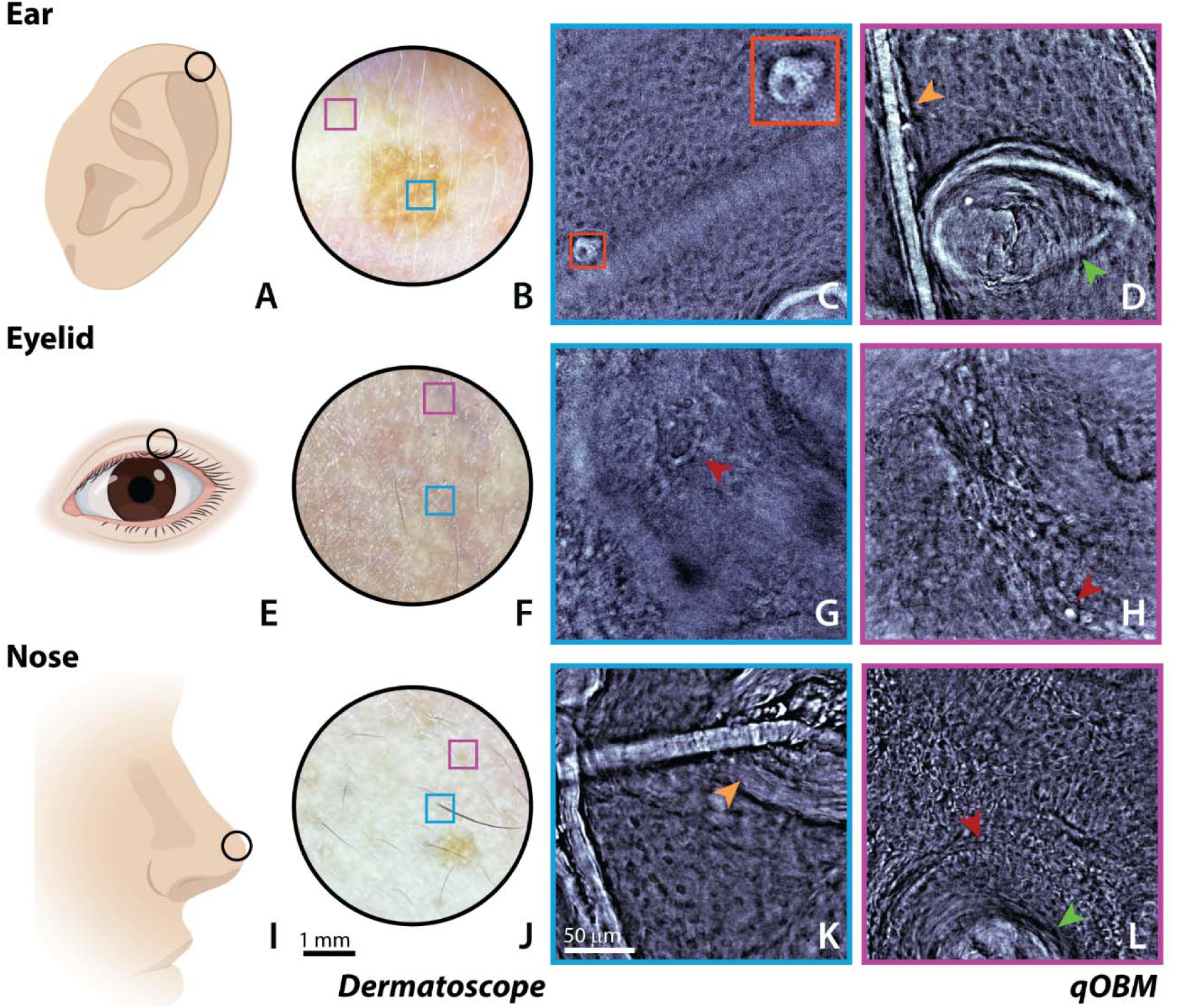
Volumetric qOBM probe images anatomically challenging facial areas. Typically inaccessible areas of the body to bulkier systems by can be readily imaged with the qOBM handheld probe. The helix of the ear (area depiction in **A**, dermatoscope image in **B**) shows potential Langerhans cells (**C**, inset), keratin buildup (**D**, green arrow), and hair (**D**, orange arrow). From the eyelid (area depiction in **E**, dermatoscope image in **F**), we visualize lateralized blood flow (red arrows in **G**, **H**). The tip of the nose (area depiction in **I,** dermatoscope image in **J**) shows hair and hair follicles (orange arrow in **K**), lateralized blood flow (red arrow in **L**), and sebaceous build up in a skin pore (green arrow in **L**).

In **Fig. 4E-H**, we show an area depiction (**Fig. 4E**), dermatoscopy (**Fig. 4F**) and qOBM (**Fig. 4G-H**) images of the eyelid. The eyelid has relatively thin epidermis,^27^ enabling visualization of the dermis. Instead of the characteristic cobblestone pattern organized keratinocytes display in the epidermis, we see the long, fibrillar strands of collagen and the amorphous extracellular matrix of the dermis. In addition, we see the lateralized blood flow present at the upper layers of the dermis (red arrows in **Fig. 4G-H**, **Supp. Videos 3-4**), a contrast to the looped capillaries of the rete ridges seen in the stratum basale layer of the epidermis.

In **Fig. 4I-L**, we show an area depiction (**Fig. 4I**), dermatoscopy (**Fig. 4J**) and qOBM (**Fig. 4K-L**) images of the tip of the nose. At more superficial layers of the epidermis, we see larger keratinocytes with lower nuclear density and part of a hair follicle (orange arrow, **Fig. 4K**). Deeper into the epidermis and dermis, we see sebaceous filament build up (green arrow, **Fig. 4L**) and more lateralized blood flow (red arrow in **Fig. 4L**, **Supp. Video 5**). The handheld probe enables monitoring in these anatomically challenging facial areas that larger, bulkier systems would struggle to image; real-world photographs of the probe imaging these areas are available upon request. In summary, the handheld qOBM probe enables *in vivo,* real time, volumetric imaging of the healthy epidermis and top layers of the dermis across all skin types and anatomical sites. These results of this section (**Figs. 2-4**) demonstrate the handheld probe highlights the dynamic 3D volumetric microstructure of the skin across all Fitzpatrick skin types, while its easy maneuverability facilitates imaging of anatomically challenging sites that can be difficult to reach with bulkier systems.

### 2.3. The handheld qOBM probe identifies histologically relevant features of healthy and diseased skin in murine melanoma model

Next, we investigate the diagnostic potential of the handheld qOBM probe by conducting a case study with a murine melanoma model. We image a 3 mm melanotic melanoma and surrounding healthy tissue first with the benchtop qOBM system (**Fig. 5A**). As the tissue transitions from healthy to disease (**Fig. 5A** and **5B**), we see a trend towards disorganization in the cellular network, nuclear atypia, and a higher nuclear to cytoplasmic (N:C) ratio, which are common indicators of tumor pathology.^33,34^ We then compare the observations made with the bench top system to those made with the handheld probe by specifically imaging healthy, transitional, and melanotic melanoma murine epidermal tissue.

**Figure 5.**
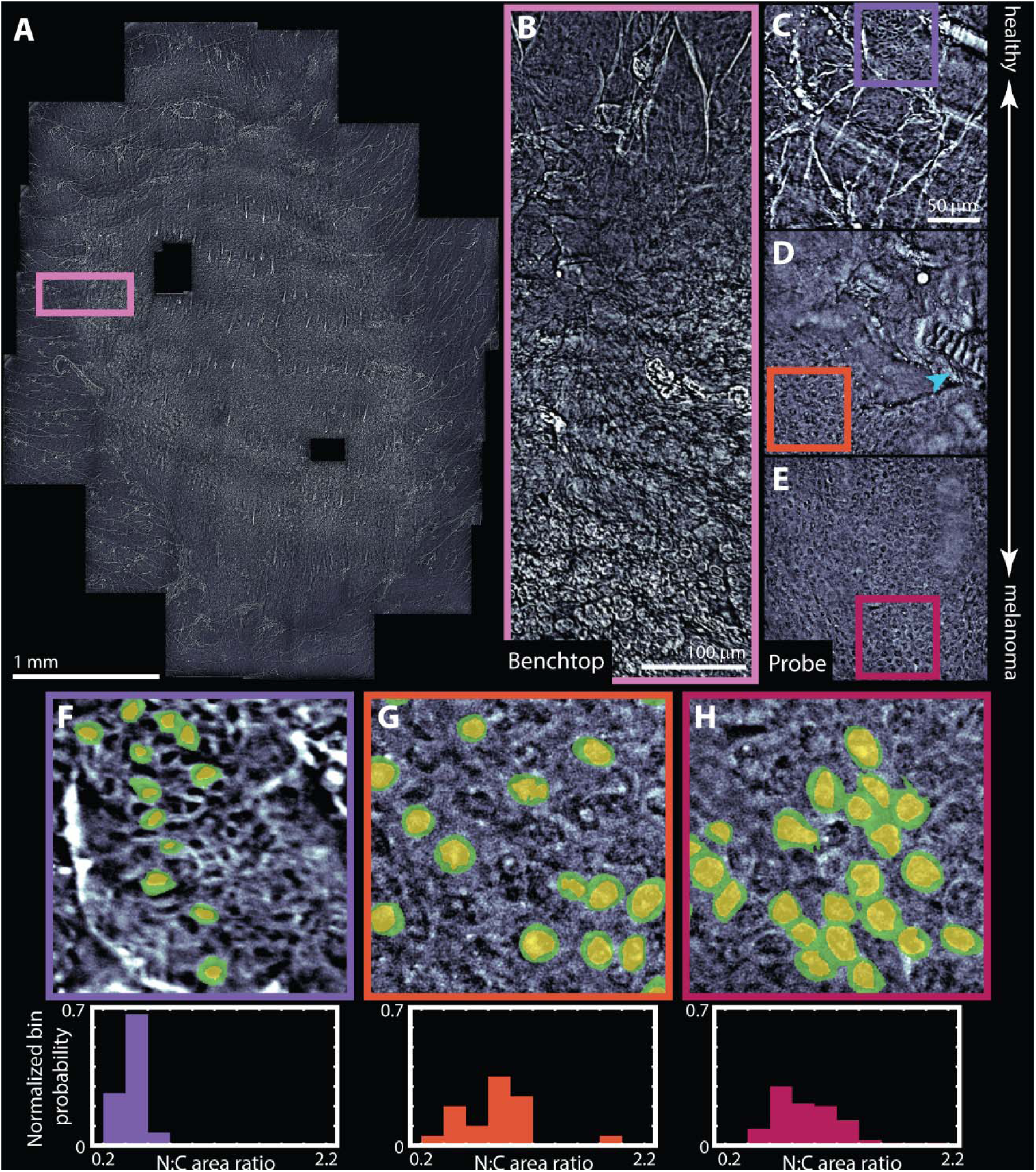
Imaging melanoma on murine models. A 3 mm mouse melanotic melanoma induced in a murine model was imaged first with the benchtop qOBM system (**A**) to observe the transition from healthy to melanoma tissue along the lesion (**B**). Regions of healthy (**C**), transitional (**D**), and melanoma (**E**) tissue were corroborated with the handheld qOBM probe. (**F**), (**G**), and (**H**) demonstrate insets highlighting nuclear (yellow shading) and cytoplasmic (green shading) regions from healthy (**C**), transitional (**D**), and tumor (**E**) skin. Corresponding distributions of calculated nuclear to cytoplasmic (N:C) area ratio for representative cells in each image are shown below the insets. Insets are 75 µm x 75 µm.

Healthy regions of the skin (**Fig. 5C**) imaged with the handheld probe show cobblestoning indicating organized epithelia and regular striations indicative of skin microrelief; we also observe a regular nuclear size and N:C ratio, (purple inset, **Fig. 5F**). It is important to note that healthy murine epidermis is exceedingly thin (<30 um) and thus dominated by skin microrelief (unlike thicker human epidermis, which demonstrates this microrelief most prominently in the superficial stratum corneum).^35^ The ability of the probe to vary the imaging depth to the precise depth of interest is critical here and could not have been achieved with our prior first-generation probe which had a fixed focal length. As the probe is transitioned to the melanotic melanoma tissue, we see some erosion of the thin epidermis, evidenced by the presence of a disrupted hair follicle (green arrow, **Fig. 5D**). As we move completely into the tumor region with the probe (**Fig. 5E**), the keratinocytes are clearly larger, have a high N:C (**Fig. 5H**) and show signs of greater disorganization without any skin microrelief, potentially indicating epidermal thickening and greater rates of cellular proliferation.

We perform a simple quantitative comparison of the N:C ratios between cells found in the images of normal, transitioning, and tumor skin. From a representative sample of cells in each image, we segment the nuclear (highlighted in yellow in **Figs. 5F-H**) and cytoplasmic areas (highlighted in green in **Figs. 5F-H**) and calculate the N:C ratio for keratinocytes in normal, transitioning, and tumor skin. From these representative N:C distributions, normal keratinocytes exhibit a smaller and more uniform N:C ratios in comparison to the transitional and tumor keratinocytes. These findings corroborate the subtle visual changes observed in the transition from healthy to tumor tissue. Not only do these preliminary findings reveal disease-associated changes in murine skin microstructure, but they also demonstrate the probe’s potential to distinguish these changes through direct visualization and quantification methods. Although further validation is required to establish diagnostic utility, they motivate future studies to establish the histopathological significance of observed features and the diagnostic potential of this technique.

### 2.4. The handheld qOBM probe comprehensively assesses human nevi in vivo

Integrating the handheld probe into a dermatology clinic presents its own set of inherent challenges that cannot be readily controlled in a clinical setting.To validate the handheld probe’s usage for skin lesions in humans, we demonstrate two nevi imaged with dermoscopy and the handheld probe in a dermatology clinic setting, specifically Emory Univeristy Hospital (**Figure 6**). All human studies were approved by the IRB (Protocol # STUDY0008502) and informed consent was obtained. Our first patient presented with a dysplastic nevus on their lower back. **Fig. 6A** shows a dermoscopic image encompassing both the lesion and the surrounding healthy skin, and **Figs. 6B** and **6C** show qOBM images of the lesion and adjacent healthy skin, respectively. Both qOBM images were captured around the stratum basale layer of the epidermis. Here, the dysplastic nevus shows random, irregular bright spots (orange arrows, **Fig. 6B**) not present in the surrounding regions of the healthy skin (**Fig. 6C**), indicative of disorganized melanocytes. Also, the overall pattern of the skin is less organized; in addition, the image is brighter overall indicative of a higher refractive index composition, likely from the higher amount of pigment. To capture this diagnostically relevant information in a living, breathing patient within the short timeframe of a clinical visit can be exceedingly challenging. For example, the natural breathing pattern of the patient can implicate capturing the desired imaging plane for a sustained period time. The volumetric capabilities of the handheld qOBM probe can mitigate this disruption, as the slight axial shifts from regular breathing can be compensated for by ETL tuning. For the dysplastic nevus, we reactively tune over an axial range of 38 µm to find the precise plane of interest; for the normal skin, we reactively tune over an axial range of 46 µm to match the approximate epidermal layer found in the dysplastic nevus for direct comparison. We demonstrate the process towards finding the desired plane of interest in the dysplastic nevus and normal skin for this patient in **Supplementary Videos 6** and **7** respectively.

**Figure 6.**
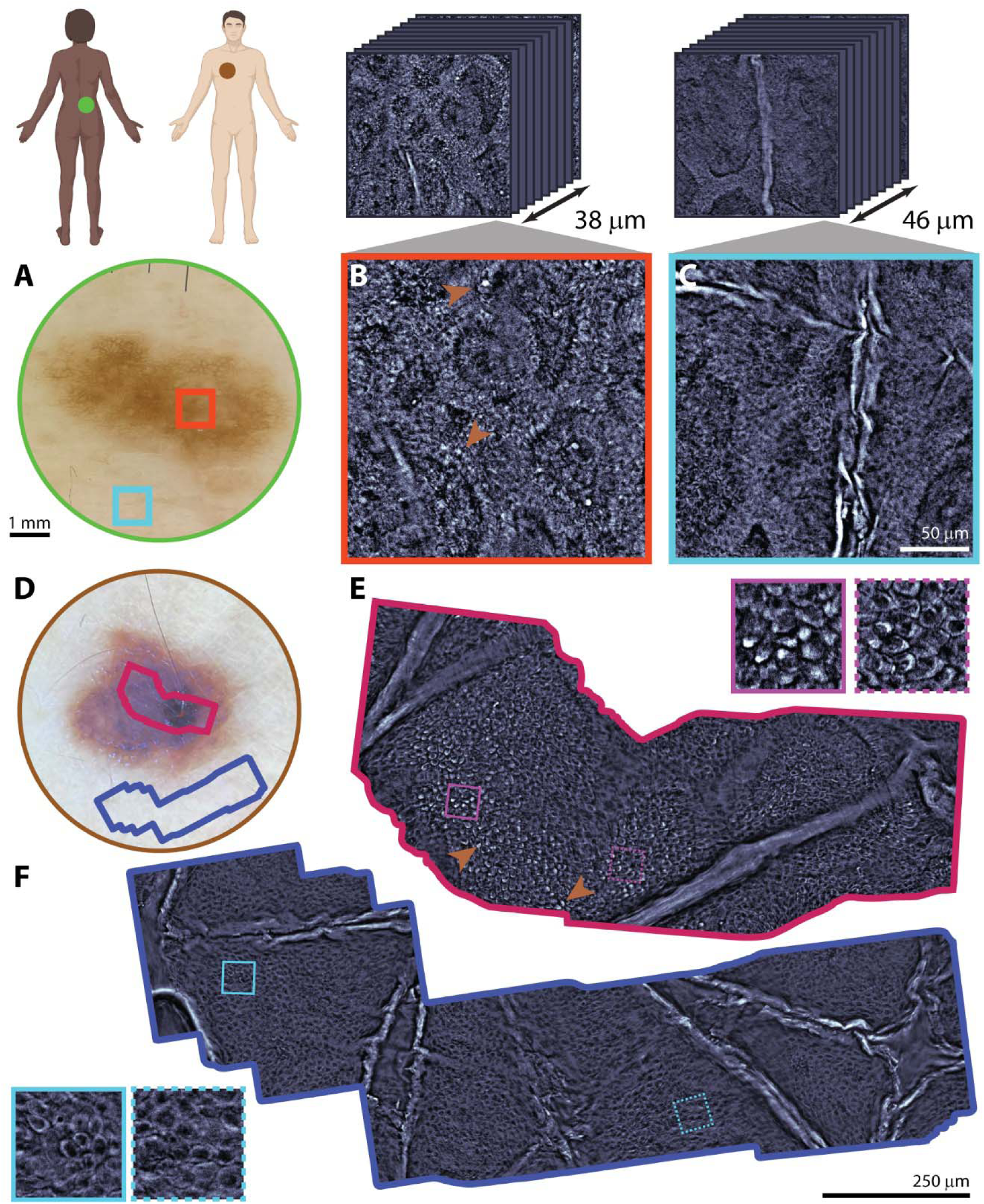
Imaging human nevi. A dysplastic nevus found on the lower back (dermatoscope image in **A**) is shown. Probe images of abnormal regions of the nevus (**B**) show random highly melanocytic cells (orange arrows) whereas normal regions of the nevus (**C**) do not display this random distribution of melanocytes. The axial scanning range used to locate the displayed region of the dysplastic nevus in (**B**) in the patient was 38 µm and was 46 µm for the normal region in (**C**). A melanocytic compound nevus found on the upper chest (dermatoscope image in **D**) is shown. Areas with higher (**D**) and lower (**E**) melanin content were stitched together from individual probe images. Regions with a high density of melanocytic cells are found in the melanotic region of the lesion (**E**, insets), whereas areas external to the lesion displays typically uniform keratinocyte morphology (**F**, insets).

The second patient displayed a highly melanotic benign compound nevus (dermoscopy in **Fig. 6D**). Here we stitched ∼1mm FOVs of the lesion and surrounding skin at the stratum spinosum layer of the epidermis (**Figs. 6E** and **6F**, respectively). The qOBM images of the nevus show patches of bright melanocytes (orange arrows, solid line inset **Fig. 6E**) clustered into melanocytic nests with a range of cellular morphology (dashed line inset **Fig. 6E**), while in the surrounding normal skin typical keratinocytic morphology (insets **Fig. 6F**) and a lack of melanocytes are seen. This larger field of view enables the viewer to visualize melanocytes clustering/nesting among typical keratinocytes. Real-time acquisition of the area stitched in **Figs. 6E-F** can seen in **Supplementary Videos 8** and **9** respectively. Although this nevus is benign, it does exhibit some of the ABCDE criteria used for initial visual diagnosis of cutaneous melanoma.^36^ It is asymmetric, has an irregular border, and has some color variation which a different dermatologist (with less dermatoscopy experience) may find suspecious enough to biopsy. However, the presence of this small melanocytic nest within the larger expanse of healthy keratinocytes without cellular atypia supports its diagnosis with more certainty as a benign nevus.^37^ Without the easy manueverability and integratable stitching capabilities compatible with the qOBM handheld probe, melanocyte clusters—if viewed in isolation—could potentially cause the lesion to appear suspicious. Thus, the handheld probe’s volumetric imaging capabilities and maneuverability not only enable dynamic visualization of skin microstructure in anatomically challenging regions but also facilitate precise targeting of regions of interest of varying sizes in variable clinical settings.

## 3. CONCLUSION

In this work, we presented a highly maneuverable, handheld probe capable of providing high-resolution, 3D visualization of the skin microstructure. The volumetric handheld probe integrates the complementary strengths of existing skin imaging techniques, combining dermatoscopy’s maneuverability with the high-resolution, volumetric visualization of skin microstructure of RCM and OCT. As shown in healthy human skin, our probe is capable of visualizing the dynamic, layered microstructure of the epidermis and the top of the dermis across all Fitzpatrick skin types. The easy manueverability of the probe enables imaging in anatomically challenging areas typically hard to reach for bulkier systems, such as the ear, eyelid, and tip of the nose. We presented additional case studies that further illustrate the potential clinical applications of the volumetric handheld probe. Using an *ex vivo* murine model of melanoma, we demonstrated visual and quantifiable alterations in skin microstructure associated with melanoma, highlighting the probe’s potential to identify image-based features that distinguish tumor from healthy skin; these findings motivate further studies validating this probe’s diagnostic potential. We also demonstrated the utility of the probe design in the clinical setting while assessing human nevi *in vivo*; the probe’s volumetric capabilities and easy maneuverability enable precise localization of regions of interest.

The central strength of the volumetric qOBM imaging probe is that it combines the handheld, compact, low cost characteristics of the widely-used dermatoscope and the high-resolution, volumetric information of RCM and OCT to provide subcellular information at the dermatologist’s bedside or other point of care settings in real time. Because of the clear histological information provided, this probe could potentially inform diagnosis from anatomic sites across the body by pathology interpretation (and potentially automated computational methods) better than visualization by low-resolution dermatoscopy. The probe is a portable technology that can be used anywhere and potentially integrated with other devices, such as existing technologies used for bedside dermatological evaluation. In addition to being low cost, there are also no moving components in the handheld system, making it robust. qOBM itself is an adaptable technique that can be converted into a larger benchtop system for high-resolution, large FOV evaluation with volumetric scanning,^19^ a smaller system for even greater locational flexibility,^38^ or a multimodal system capable of visualizing complementary contrast mechanisms, such as fluorescence.^39–41^

Like many optical imaging techniques, qOBM’s penetration depth is limited to 200-300 µm at the wavelengths used in this work (850nm). However, with longer wavelengths (e.g., 1300 nm), we expect to be able to image deeper into the tissue, as demonstrated by a scanning benchtop qOBM system that enables >0.5 mm imaging depths.^42^ *In vivo*-compatible optical clearing agents, such as tartrazine and fluorescein, have also enhanced the depth penetration of qOBM to ∼0.5 mm with 850 nm illumination and up to 1.7 mm with 1300 nm illumination with subcellular resolution.^30,43^ *In vivo* compatible clearing agents are expected to be promising imaging aids once approved for human *in vivo* use. Like any handheld imaging technique, the qOBM probe has its share of motion artifacts, which can prevent precise monitoring of functional information such as blood flow and decreases image quality with motion blur. This current version of the probe utilizes four captures to create a single qOBM image, which enables imaging at a quarter of the camera acquisition rate. We have recently developed single-capture qOBM (SCqOBM) which generates a qOBM image from a single capture instead of four, thereby enabling qOBM imaging at the camera rate.^44^ Faster imaging will improve real-time functional visualization of blood flow and probe-roving image quality. Another limitation is that the current imaging area of the probe is small (∼350 µm x 350 µm). However, with the additional stitching capabilities illustrated, we can use the handheld probe to perform real-time video-mosaicking as previously accomplished with RCM techniques.^45^

In the future, we aim for the probe to act as a companion for existing imaging techniques, such as the dermatoscope and RCM/OCT, and improve diagnostic assessment by providing real-time histopathological information at the bedside. The potential cases for clinical use are numerous. Cancer screening is a clear clinical use case, but monitoring the treatment and immunological response of cancer sites during/after sequential radiation treatment is also a promising avenue. The probe could be used as an adjunct device during Moh’s surgery by providing a noninvasive optical biopsy of skin surrounding lesions after excision to ensure complete resection. In summary, this work fills the gap in skin imaging techiques between low-resolution, highly maneuverable systems like dermatoscopy and high-resolution, bulky systems like OCT and RCM. By filling this gap, the volumetric handheld probe has the potential to provide dermatologists with pathology-like insights directly at the point of care, informing clinical assessment and treatment decisions across a range of care environments. As the body’s first line of defense against environmental threats, the skin warrants comprehensive imaging approaches capable of capturing its complex structure and supporting clinical evaluation across diverse clinical settings.

## 4. MATERIALS AND METHODS

*Murine melanoma model:* In this work, intradermal melanoma murine models were used for melanoma imaging.^46^ B16F10 murine melanoma cells were cultured under sterile conditions in DMEM containing 10% FBS and 1% penicillin-streptomycin-amphotericin B and were collected using 0.05% trypsin-EDTA. To inoculate B16F10 tumors, the dorsal region of female C56BL/6 mice was shaved and 100K B16F10 cells were injected intradermally in sterile saline while the mice were under isoflurane anesthesia. Normal and tumor-containing murine skin from seven mice were imaged 6-7 days after intradermal tumoral injection with both the benchtop and probe qOBM systems. For this initial evaluation we only imaged four mice with the probe. Mice were imaged at two positions – at the intradermal tumor site along the dorsal thoracic area or on the abdomen of the mouse and at a normal skin site, often adjacent to the tumor or at the abdomen. Prior to imaging, the mouse underwent a two-step hair removal process at the imaging ROI: initial shaving with an electric razor and subsequent 5-minute application and removal of NAIR hair removal lotion. Further details about the IACUC-approved protocol employed when handling the mouse can be found in prior work.^46^

*Handheld probe image stitching methodology:* To create panoramas from the videos captured by the handheld qOBM probe, we developed a custom feature-based stitching pipeline using Scale-Invariant Feature Transform (SIFT) features.^47,48^ Random Sample Consensus (RANSAC) was used to reject outliers and blending was performed via a raw pasting method and a gradient-weighted blending method.^49^ An in-depth description of the methodology behind this stitching algorithm will be published in a forthcoming paper. The algorithm is loosely based on video mosaicking methods developed for RCM^45^ but tailored specifically for qOBM image stitching.

*Cellular and nuclear segmentation for nuclear to cytoplasmic ratio calculations:* Initial cellular segmentation was performed with Cellpose^50^ integration with Matlab 2023b. After cellular segmentation, nuclei were manually segmented within the cellular area; incorrectly segmented cells were discarded from the analysis. The cytoplasmic area was calculated as the nuclear area subtracted from the total cell area. The nuclear to cytoplasmic (N:C) area ratio was calculated as the nuclear area divided by the cytoplasmic area.

## ACKNOWLEDGEMENTS

The authors would like to acknowledge the healthy volunteers for imaging: RG, RL, JL, GC, and VG. The authors would like to acknowledge funding from the ARCS Foundation, the Georgia Clinical and Translational Science Alliance; the National Institute of General Medical Sciences (R35GM147437); Georgia Institute of Technology; St Jude Children’s Research Hospital.

## DATA AVAILABILITY STATEMENT

The data supporting the findings of this study are available from the corresponding author upon reasonable request. Data include representative qOBM image datasets supporting the development and characterization of the handheld volumetric qOBM probe and its demonstration in skin. Human imaging data are subject to applicable ethical and privacy restrictions and may be shared only where permitted by the relevant approvals and consent.

## Funding

The authors would like to acknowledge funding from the ARCS Foundation, the Georgia Clinical and Translational Science Alliance; the National Institute of General Medical Sciences (R35GM147437); Georgia Institute of Technology; St Jude Children’s Research Hospital.

## Notes

### Competing Interest Statement

The authors have declared no competing interest.

### Author Declarations

IRB of Emory University gave ethical approval for this work.

