## Supplementary Information for "Handheld, quantitative phase imaging probe for skin evaluation at subcellular resolution *in vivo*, in real-time, and in 3D"

### Supplemental Information

#### 1 Volumetric qOBM probe axial range derivation

Here we describe both the theoretical derivation of the probe's axial range with the paraxial approximation and its validation in experimental practice. We complete this scenario for two cases – (1) the ETL acting on the converging light as the image is relayed from the tube lens to the camera and (2) the ETL acting on the light in infinity-space, after passing though the objective.

##### 1.1 Basic theoretical approach for deriving axial range

With the paraxial approximation, any given system can be defined as:

$$\begin{pmatrix} y_f \\ \alpha_f \end{pmatrix} = \begin{pmatrix} A & B \\ C & D \end{pmatrix} \cdot \begin{pmatrix} y_i \\ \alpha_i \end{pmatrix}$$

where $y_i$ and $\alpha_i$ represent the incident position perpendicular to the optical axis and angle of the beam and $y_f$ and $\alpha_f$ represent the final position and angle of the beam, respectively. The $ABCD$ matrix here describes the optical system the rays travels through.

We can think about our object as a single point source along the optical axis. This object has a singular position $y_i = 0$ with rays emanating at angles, $\alpha_i$. For this point to be in focus, all of these rays must converge to $y_f = 0$ at the imaging plane, regardless of their final beam angle ($\alpha_f$). From the paraxial approximation, we can relate $y_i$ and $y_f$:

$$\begin{aligned} y_f &= A \cdot y_i + B \cdot \alpha_i \\ 0 &= A \cdot 0 + B \cdot \alpha_i \\ 0 &= B \cdot \alpha_i \end{aligned}$$

Because all rays (except for one) will have non-zero initial and final angles, $B = 0$ for the point source to be in focus at the image plane.

The volumetric probe system can also be modeled with the paraxial approximation, such that the entire system $ABCD$ matrix ($M$) converges to where $B = 0$. We model two different cases of the volumetric probe configuration.

##### 1.2 Case 1: ETL placed behind tube lens

When the electronically tunable lens (**ETL**) is placed between the tube lens and the camera (see **Fig. 1**), it acts on the converging beam created by the tube lens.

###### 1.2.1 Theoretical derivation with the paraxial approximation of Case 1

We can model this system from object plane to image plane with ray transfer matrices. Here we use $P$ for the propagation matrix, $N$ for refraction at a flat surface, and $L$ for matrix of a lens (detailed definitions of the matrices and derivations for the thick lens approximations used here

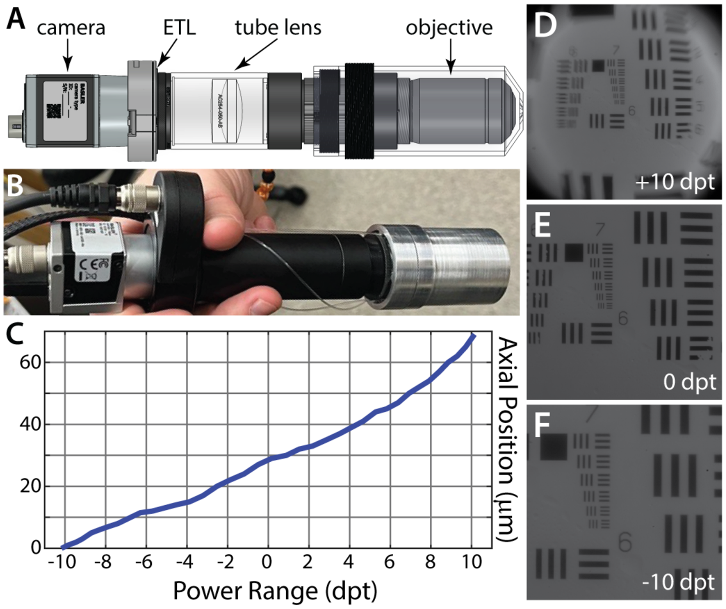


Figure 1: **Case 1 system configuration.** For Case 1, the ETL is placed between the camera and the tube lens as seen in CAD design (**A**) and in physical implementation (**B**). Sequential, in-focus images of the 1951 USAF resolution target were taken throughout the diopter range (examples at +10 dpt (**D**), 0 dpt (**E**), and -10 dpt (**F**)), and the axial position was noted. The full axial range over the power spectrum of the ETL is shown in **C**.

can be found in the Appendix (**Sections 5.1-5.2**)). If we model the system where the sample is a glass target in air:

$$\begin{aligned} M =& P(x_{CE}) \cdot L(ETL_D) \cdot P(f_{TL} - x_{CE} - x_{ETL}) \cdot L(TL) \cdot \\ & P(x_I) \cdot L_{thin}(f_{obj}) \cdot P(x_{WD}) \cdot \\ & N(n_{glass}, n_{air}) \cdot P(x_{CG}) \cdot N(n_{air}, n_{glass}) \cdot P(x_{sample}) \end{aligned}$$

where $x_{CE}$ is the distance between the camera and the ETL, $L(ETL_D)$ is the ETL modeled as a thick lens (see **Section 5.2.2**) at a power of $D$ diopters, $f_{TL}$ is the focal length of the tube lens, $x_{ETL}$ is the thickness of the ETL, $L(TL)$ is the tube lens modeled as a thick lens (see **Section 5.2.1**), $x_I$ is the length between the objective and the tube lens, $L_{thin}(f_{obj})$ is the objective modeled as a thin lens with effective focal length $f_{obj}$, $x_{WD}$ is the working distance of the objective, $n_{glass}$ is the refractive index of glass, $n_{air}$ is the refractive index of air, $x_{CG}$ is the thickness of the cover glass, and $x_{sample}$ is the remaining distance to the target.

All of these values are known and fixed for the system except for $x_{sample}$ which depends on the variable ETL focusing power, $ETL_D$. Thus, the system matrix can be simplified to:

$$M = F(ETL_D) \cdot P(x_{sample})$$
$$\begin{pmatrix} A & B \\ C & D \end{pmatrix} = F(ETL_D) \cdot P(x_{sample})$$

where $B = 0$, and $F$ is all terms in system matrix except for the propagation from the sample to the cover glass. We can solve this matrix equation for $x_{sample}$ at both ends of the ETL's diopter range. When the ETL is positioned 19 mm from the camera, for ETL configuration at

+10 diopters, we arrive at $x_{sample} = 417\mu m$. For ETL configuration at -10 diopters, we arrive at $x_{sample} = 481\mu m$, which gives an axial range of $64\mu m$ for this particular system configuration. The theoretical magnification ranges from $8.6 - 16.2X$. When simulating in tissue, our axial range extends to $89\mu m$ with the same magnification range.

##### 1.2.2 Experimental validation of Case 1

To experimentally validate the axial range calculated from the paraxial approximation, we align the system to its focus throughout the diopter range of system. We used a 1951 USAF resolution target to determine the focal point at each diopter value and measured the axial range with a manually positioned z-stage micrometer; images of the target at selected diopter values are shown in **Fig. 1D-F**. The full axial range (**Fig. 1C**) for this configuration along the ETL power range was experimentally determined to be $69\mu m$, and the magnification ranged from $10.2 - 21.2X$. The axial range matches closely to the theoretically derived range of $64\mu m$ shown in **Section 1.2.1**. With this configuration, small changes in the theoretically reported distance between the camera and ETL can vastly alter the axial range, so slight measurement errors may contribute to slight discrepancy in axial range. The theoretical and experimentally-derived magnification ranges differ by about 20%, but this discrepancy can be explained by the simplification of the lens modeling of the objective.

#### 1.3 Case 2: ETL placed behind objective

When the ETL is placed between the objective and the tube (see **Fig. 2**), it acts on the beam created by the objective.

##### 1.3.1 Theoretical derivation with the paraxial approximation of Case 2

We can model this system from object plane to image plane with ray transfer matrices (note definitions of the types of ray transfer matrices and derivations for the thick lens approximations used here in the Appendix (**Sections 5.1-5.2**)). If we model the system where the sample is a glass target in air:

$$\begin{aligned} M = &P(f_{TL}) \cdot L(TL) \cdot \\ &P(x_I - x_{EO}) \cdot L(ETL_D) \cdot P(x_{EO}) \cdot \\ &L_{thin}(f_{obj}) \cdot P(x_{WD}) \cdot \\ &N(n_{glass}, n_{air}) \cdot P(x_{CG}) \cdot N(n_{air}, n_{glass}) \cdot P(x_{sample}) \end{aligned}$$

where $x_{EO}$ is the distance between the objective and the ETL and all other definitions remain the same as described in **Section 1.2.1**.

Following the same procedure as noted in **Section 1.2.1**, we can solve this matrix equation for $x_{sample}$ at both ends of the ETL's focusing power range. When the ETL is positioned 38.9 mm from the objective, for ETL configuration at +10 diopters, we arrive at $x_{sample} = 269\mu m$. For ETL configuration at -10 diopters, we arrive at $x_{sample} = 581\mu m$, which gives an axial range of $312\mu m$ for this particular system configuration. The theoretical magnification range for this system is $7.7x$ to $20.9x$. When simulating in tissue, our axial range extends to $436\mu m$ with the same magnification range.

##### 1.3.2 Experimental validation of of Case 2

To experimentally validate the axial range calculated from the paraxial approximation for Case 2, we align the system to its focus throughout the diopter range of system. We used a 1951 USAF resolution target to determine the focal point at each diopter value and measured the axial range with a manually positioned z-stage micrometer; images of the target at selected diopter values are shown in **Fig. 2D-F**. The full axial range (**Fig. 2C**) for this configuration along the ETL power range was experimentally determined to be $313\mu m$, and the magnification ranged from $6.5 - 23.4X$. Similar to the results from Case 1, the axial range matches closely to the theoretically derived range of $312\mu m$ shown in **Section 1.3.1**. As was reported with Case 1, the theoretical and experimentally-derived magnification ranges differ by less than 20%. Again, this discrepancy can be explained by the simplification of the lens modeling of the objective.

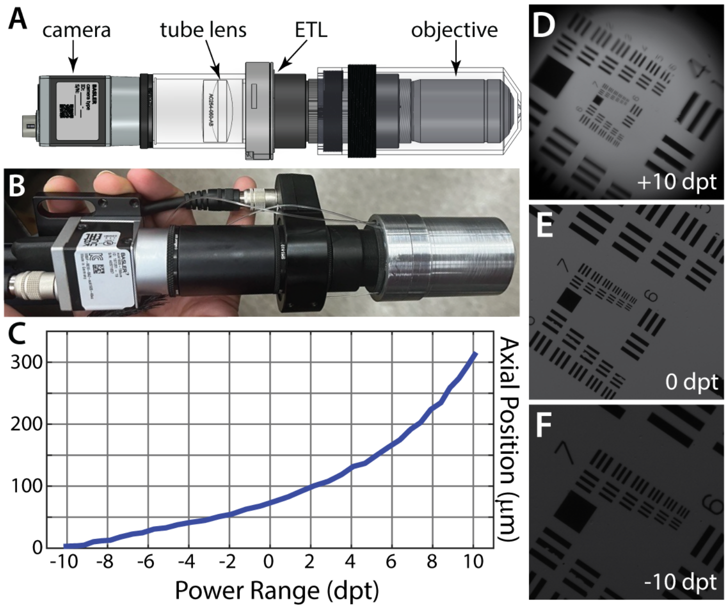


Figure 2: **Case 2 system configuration.** For Case 2, the ETL is placed between the tube lens and the objective as seen in CAD design (**A**) and in physical implementation (**B**). Sequential, in-focus images of the 1951 USAF resolution target were taken throughout the diopter range (examples at +10 dpt (**D**), 0 dpt (**E**), and -10 dpt (**F**)), and the axial position was noted. The full axial range over the power spectrum of the ETL is shown in **C**.

#### 2 Monte Carlo Simulations for Optimal Illumination Placement

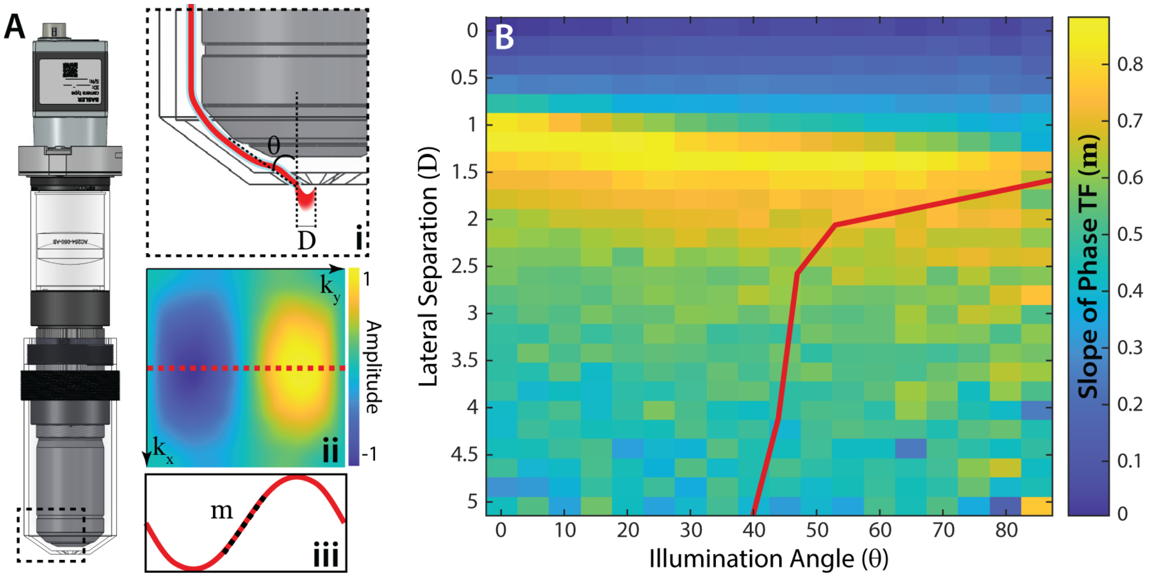


Figure 3: **Optimizing phase sensitivity with Monte Carlo simulations.** Through Monte Carlo simulations, we determine the optimal placement of the multimodal fiber used for illumination for the volumetric probe (**A**). The multimode fiber illumination is placed at lateral separation $D$ from the center of the optical axis and at an angle $\theta$, shown in inset **Ai**. The optical transfer function of the system (**Aii**) is projected through one axis of frequency space (**Aiii**). The slope of this projection is calculated across a range of lateral separations and angles (**B**) as a proxy for phase sensitivity of the system, where all configurations to the right of the red line are physically possible.

Monte Carlo simulations were run to optimize placement of the illuminating multimodal fibers (MMFs). The lateral position $D$ and angle $\theta$ of the MMF (see **Fig. 3Ai**) can be optimized to maximize the phase sensitivity of the system. A proxy for the phase sensitivity is the slope of the optical transfer function (TF), as shown in **Fig. 3Aii**. The TF is projected across one dimension (red lines in **Fig. 3Aii-iii**) and the slope at the center of the projection is calculated (dotted line **Fig. 3Aiii**). A map of these slopes of the TF through a range of lateral separations $(0 < D < 5mm)$ and illumination angles $(0 < \theta < 85°)$ is shown in **Fig. 3B**. The optimal position where the slope is highest occurs when $D = 1.2mm$ and $\theta = 15°$. Due to the diameter of the MMFs, this configuration is impossible. A line delineating physically possible configurations is indicated is shown in **Fig. 3B**; all configurations to the right of the line are physically possible. For our current configuration, we have selected a physically plausible position with moderate phase sensitivity, where $D = 2.54mm$ and $\theta = 60°$. Additional documentation detailing this optimization process for qOBM systems can be found in prior work.[1]

#### 3 Image Entropy Across Fitzpatrick Skin Type

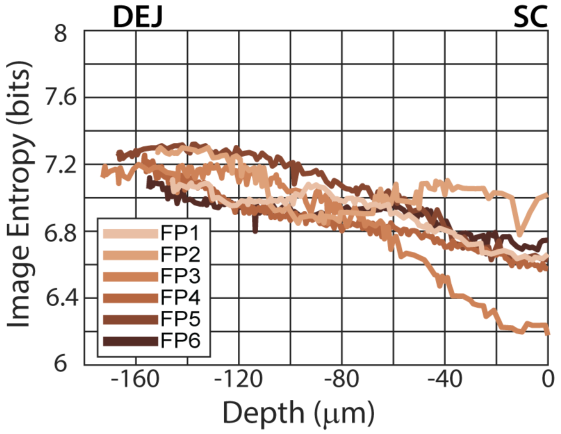


Figure 4: Whole image entropy plotted over increasing depth ($\mu m$) over epidermal tissue for six volumes across the Fitzpatrick (FP) scale shown in **Fig. 3** and **Supplementary Video 2**. The most superficial layer of the epidermis, the stratum corneum (SC), is set to 0 $\mu m$ for all subjects. With variable epidermal thicknesses among subjects, the dermal epidermal junction (DEJ) is at different depths.

We plot the whole image entropy across the epidermal imaging depth for the six different Fitpatrick type volumes shown in **Fig. 3** in the main text and **Supplementary Video 2**. Entropy measurements start at the surface (stratum corneum, set to 0 $\mu m$) and continue to the dermal-epidermal junction (DEJ). The general increase in entropy with depth is similar across all Fitzpatrick types, further corroborating the volumetric probe's ability to image all skin types. Lower entropies for the superficial layer are indicative of some oversaturation of the images at the stratum corneum; this oversaturation is particularly prominent in the superficial layers of the FP3 volunteer. Note the natural variability in epidermal thickness as the final DEJ position, ranging from -150-170 $\mu m$, differs among subjects.

#### 4 Extraneous Supplemental Media

Here we display or describe extraneous media referenced by main manuscript:

**Supplementary Video 1.** 3D visualization of volumetric imaging of the whole epidermis shown in **Fig. 2**. Scale bar 50$\mu m$, frame rate 10 frames per second (fps).
**Supplementary Video 2.** 3D visualization of volumetric imaging of the epidermis of subjects from all six Fitzpatrick (FP) skin tones shown in **Fig. 3**. To ensure video synchronization, these videos cover the same diopter range. The most superficial layer of the epidermis, the stratum corneum (SC), comes into focus at different axial depths within the videos. For FP1-6, the SC comes into focus at 53 $\mu m$, 50 $\mu m$, 30 $\mu m$, 46 $\mu m$, 34 $\mu m$, 48 $\mu m$, respectively. Scale bar 50$\mu m$, frame rate 10 fps.
**Supplementary Video 3.** Roving, real-time visualization of eyelid imaging shown in **Fig. 4G**. Scale bar 50$\mu m$, frame rate 10 fps.
**Supplementary Video 4.** Roving, real-time visualization of eyelid imaging shown in **Fig. 4H**. Scale bar 50$\mu m$, frame rate 10 fps.

**Supplementary Video 5.** Roving, real-time visualization of tip of nose imaging shown in **Fig. 4K**. Scale bar 50$\mu m$, frame rate 10 fps.
**Supplementary Video 6.** Real-time axial range tuning to identify appropriate plane for dysplastic nevus shown in **Fig. 6B**. Scale bar 50$\mu m$, frame rate 10 fps.
**Supplementary Video 7.** Real-time axial range tuning to identify appropriate plane for dysplastic nevus shown in **Fig. 6C**. Scale bar 50$\mu m$, frame rate 10 fps.
**Supplementary Video 8.** Real-time acquisition of images stitched in **Fig. 6E** mosaic. Scale bar 50$\mu m$, frame rate 15 fps.
**Supplementary Video 9.** Real-time acquisition of images stitched in **Fig. 6F** mosaic. Scale bar 50$\mu m$, frame rate 15 fps.

### 5 Appendix

#### 5.1 Ray Transfer Matrix Definitions with ABCD Matrices

Here we describe the typical ray transfer matrices used in the paraxial approximation of the system. If we model the whole system using typical ray transfer matrices, we define the propagation matrix $P(d)$:

$$P(d) = \begin{pmatrix} 1 & d \\ 0 & 1 \end{pmatrix}$$

where $d$ is the propagation distance along the optical axis (in mm) in air, and $\frac{d}{n}$ is the propagation distance along the optical axis (in mm) in a medium of refractive index $n$. We also define the thin lens matrix $L_{thin}(f)$:

$$L_{thin}(f) = \begin{pmatrix} 1 & 0 \\ -\frac{1}{f} & 1 \end{pmatrix}$$

where $f$ is the focal length (in mm) of a thin lens. We define the refractive index transition matrix $N(n_1, n_2)$ at a flat surface:

$$N(n_1, n_2) = \begin{pmatrix} 1 & 0 \\ 1 & \frac{n_1}{n_2} \end{pmatrix}$$

where $n1$ is refractive index of the initial medium and $n2$ is the refractive index of the subsequent medium. Similarly, we define the refractive index transition matrix $R(n_1, n_2, r)$ at a curved surface:

$$R(n_1, n_2, r) = \begin{pmatrix} 1 & 0 \\ \frac{n_1 - n_2}{rn_2} & \frac{n_1}{n_2} \end{pmatrix}$$

where $n1$ is refractive index of the initial medium, $n2$ is the refractive index of the subsequent medium, and $r$ is the radius of curvature of the surface.

#### 5.2 Thick Lens Approximations

For many of the elements deployed in this design, we derive ray matrix approximations that will be employed in the larger system matrix.

##### 5.2.1 Achromatic Doublet

The tube lens used in this system is ThorLabs' f50-mm 1-inch achromatic doublet AC254-050-AB, comprised of a crown and flint as shown in **Fig. 5**. The doublet can be configured as a series of matrices:

$$\begin{aligned} L(TL) = & R(n_{flint}, n_{air}, -137.5) \cdot P(3.5) \cdot \\ & R(n_{crown}, n_{flint}, -28.8) \cdot P(9.0) \cdot \\ & R(n_{air}, n_{crown}, +34.9) \end{aligned}$$

$$L(TL) = \begin{pmatrix} 0.8681 & 7.4364 \\ -0.020 & 0.9809 \end{pmatrix}$$

which confirms the focal length of this lens as 50 mm.

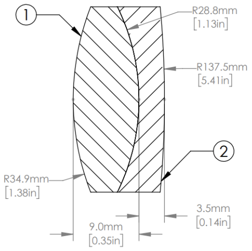


Figure 5: Schematic of Thorlabs 1-inch achromatic doublet AC254-050-AB.

###### 5.2.2 Electronically Tunable Lens (ETL)

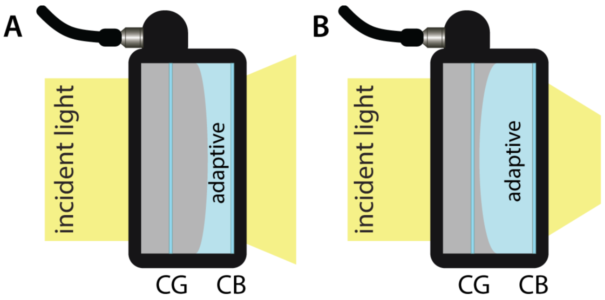


Figure 6: Drawn configurations of the ETL at (**A**) +10 diopters and (**B**) -10 diopters. The ETL is comprised of a cover glass (CG) and container backing (CB) that holds the adaptive fluid used to change the focal length of the ETL.

The ETL used is Optotune's EL-16-40-TC-VIS-20D-C-E, which has a -10 to +10 diopter range. Variation in the physical position of the ETL can be seen in **Fig. 6**. With this configuration we can construct a series of matrices to represent the ETL:

$$\begin{aligned} L(ETL_D) = & N(n_{CB}, n_{air}) \cdot P(x_{CB}) \cdot N(n_{ad}, n_{CB}) \cdot \\ & P(x_{ad_D}) \cdot R(n_{air}, n_{ad}, r_{ad_D}) \cdot P(x_{da_D}) \cdot \\ & N(n_{CG}, n_{air}) \cdot P(x_{CG}) \cdot N(n_{air}, n_{CG}) \cdot \\ & P(x_{housing}) \end{aligned}$$

where $n_{CB}$ is the refractive index of the container backing, $n_{air}$ is the refractive index of air, $x_{CB}$ is the thickness of the container backing, $n_{ad}$ is the refractive index of the adaptive fluid, $x_{ad_D}$ is the diopter-dependent thickness of the adaptive fluid, $r_{ad_D}$ is the diopter-dependent radius of curvature of the adaptive fluid, $x_{da_D}$ is the diopter-dependent distance between the adaptive fluid and the cover glass, $n_{CG}$ is the refractive index of the cover glass, $x_{CG}$ is the thickness of the cover glass, and $x_{housing}$ is the initial thickness of the housing. From this matrix we arrive at $L(ETL_{-10D})$ and $L(ETL_{+10D})$:

$$L(ETL_{-10D}) = \begin{pmatrix} 1.0169 & 15.708 \\ 0.0102 & 1.1411 \end{pmatrix}$$

$$L(ETL_{-10D}) = \begin{pmatrix} 0.9656 & 14.521 \\ -0.0096 & 0.8918 \end{pmatrix}$$

which confirms the -10 diopter ($f = -100mm$) and +10 diopter ($f = +100mm$) range of the ETL.
